# Nicotine reduces discrimination between threat and safety in the hippocampus, nucleus accumbens and amygdala

**DOI:** 10.1101/2023.05.08.23289647

**Authors:** Madeleine Mueller, Tahmine Fadai, Jonas Rauh, Jan Haaker

**Affiliations:** University Medical Center Hamburg-Eppendorf (Germany), Department of Systems Neuroscience; University Medical Center Hamburg-Eppendorf (Germany), Department of Child-and Adolescent Psychiatry and Psychotherapy; University Medical Center Hamburg-Eppendorf (Germany), Department of Psychiatry and Psychotherapy, Psychiatry Neuroimaging Branch

## Abstract

Nicotine intake by cigarettes is linked to the maintenance and development of anxiety disorders and impairs adaptive discrimination of threat and safety in humans. Yet, it is unclear if nicotine exerts a causal pharmacological effect on the affective and neural mechanisms that underlie aversive learning. We conducted a pre-registered, pseudo-randomly and double-blinded pharmacological fMRI study to investigate the effect of acute nicotine on Fear Acquisition and Extinction in non-smokers (n=88). Our results show that nicotine administration led to decreased discrimination between threat and safety in subjective fear. Nicotine furthermore decreased differential (threat vs. safety) activation in the hippocampus, which was functionally coupled with Nucleus Accumbens and amygdala, compared to placebo controls. Additionally, nicotine led to overactivation of the ventral tegmental area. This study provides mechanistic evidence that single doses of nicotine impair neural substrates of adaptive aversive learning in line with the risk for the development of pathological anxiety.

## Introduction

A key mechanism for coping with a threatening environment is aversive learning. Adaptive aversive learning enables individuals to discriminate between what is dangerous and what is safe in their surroundings. Maladaptive learning is characterized by impaired discrimination between threats and safety, for example evident in individuals that are suffering from anxiety disorders (AD)^1^. One risk factor for maladaptive aversive learning is smoking, which might be reflected by higher smoking rates in patients suffering from AD, when compared to healthy individuals^2^. Furthermore, a prospective longitudinal study indicated that smoking in healthy individuals results in a higher risk for the onset of a panic attack^3^. But the key question is how nicotine, the psychoactive ingredient in cigarette smoke, causally interferes with aversive learning, leading to impaired discrimination between safety and danger.

One possibility is that nicotine impacts the storage of aversive learning in humans. In animals, there is evidence that nicotine affects learning and associative memory formation. On the cellular level, nicotine binds to *β*2-containing nicotinic acetylcholine receptors (nAChR)^4^ and has a direct influence on synaptic plasticity such as hippocampal long term potentiation (LTP)^5,6^. The hippocampus is important to process new and salient information about threats and stores memories that results from aversive learning^7^. Such aversive learning is commonly examined in laboratory protocols of fear conditioning. During Fear Acquisition (ACQ) a conditioned stimulus (CS+) is learned as a danger signal, based on the prediction for an aversive, unconditioned stimulus (US, e.g. an electric stimulus). Another conditioned stimulus (CS-) is learned as a safety signal and predictive for the absence of the US. Indicative of adaptive learning, subjects learn to discriminate between CS+ and CS-, reflected by differential (i.e., higher for the CS+ as compared to the CS-) conditioned responses (CR).

Chronic nicotine intake, such as smoking seems to impair adaptive discrimination of threat and safety (but see^8^, which report enhanced discrimination in SCR) in fear conditioning protocols. As such, a chronic schedule of nicotine administration in rats, and smoking in humans, has shown to decrease the discrimination between CS+ and CS-^9^. Similarly, experiments in rodents revealed that the discrimination between threat and safety (contexts) is dose-dependently impaired by acute nicotine administration^10^. Supporting this finding, acute infusion of nicotine into the dorsal hippocampus (HC) disrupted safety learning in rodents^11^. These findings are related to results in humans, showing that smokers have an impaired discrimination between threat and safe contexts, when compared to non-smokers, indicated by lower discrimination in subjective fear, US expectancy and SCR^12^. A similar impairment in smokers was found in another study, which revealed less discrimination in self-reported fear towards learned threat and safety cues, compared to non-smoking individuals^13^.

While smoking seems to impair discrimination of threat and safety, it is unclear if there is a causal pharmacological mechanism by which nicotine affects the discrimination of threat and safety. Such a pharmacological effect could aid to explain maladaptive learning that is observed in smokers. The main hypothesis of this study is that acute nicotine reduces discrimination between threat and safety during aversive learning in humans.

## Methods

### Participants

For this study 88 healthy, non-smoking participants between 18 and 40 years were recruited (power analysis in supplement). Individuals confirmed to have no diagnoses of neuropsychiatric disorders, to consume less than 15 units of alcohol per week and no illegal drugs (Table S1). Additionally subjects had to be suitable for MRI measurements. All participants were non-smokers, which was defined by not being an active smoker during the time of the data acquisition and having smoked less than 200 cigarettes during their lifetime. Two participants had to be excluded from the analysis due to false statements and one participant was excluded due to too high alcohol consumption resulting in a final sample of 85 subjects (55.8% female). Three participants only completed day 1 of the study are therefore excluded from day 2 analyses. All participants gave written, informed consent to participate and received 120 C reimbursement. The study was approved by the local ethics committee (Ethikkommission der Ärztekammer Hamburg PV 5514).

### Groups

To test the effect of acute nicotine on aversive learning in a Fear Acquisition and Extinction training protocol including a reinstatement test, we compared three groups. Group 1 (Nic1) received 1mg nicotine before ACQ on day 1 and then placebo before EXT on day 2. Accordingly group 2 (Nic2) received a placebo before ACQ on day 1 and then 1mg nicotine before the EXT on day 2. Group 3 (Pla) as the control group received placebo before both days. The assignment to each group was randomized and the administration of the drug was double-blinded. The three groups did not differ in demographic parameters that were assessed in this study, except for their STAI-T score. The STAI-T score was higher in Pla, when compared Nic1 (t(25)=3.12, p=0.005) and Nic2 (t(25)=2.48, p=0.02) (Table S1).

### Experimental Procedure

The two-day paradigm (Figure 1) consisted of a Fear Acquisition (ACQ) on the first day, followed approximately 24h later by an Extinction training (EXT) with a Return of Fear manipulation (RoF) in form of a reinstatement. Both experimental days were conducted in the fMRI scanner. Depending on the pseudo-randomly assigned group, either 1mg nicotine or 1mg placebo was administered double-blinded 15min (to reach the plasma maximum) before participants were placed in the scanner and started the experiment. The Acquisition was employed on the first day. Four trials of stimulus habituation without reinforcement were followed by 3 blocks with each 8 CS+ and 8 CS-presentations. The reinforcement rate for the CS+ was 75%. On the second day, 24 hours later, EXT and the subsequent reinstatement were executed. The EXT consisted of two blocks with each 8 CS+ and 8 CS-presentations. The reinforcement rate was now 0%. Subsequently, the reinstatement took place, where four electric stimuli were presented without any context or cue information. Participants saw a black screen. That was followed by a reinstatement test with one block with 8 CS+ and 8 CS-presentations without presentation of an US (reinforcement rate=0%). Return of Fear results can be found in the Supplement. During ACQ participants saw context A and during EXT participants saw context B. In the reinstatement test, participants saw a mixture of context A and context B. This study was preregistered at the German Clinical Trials Register (Deutsches Register Klinischer Studien (DRKS); DRKS-ID: DRKS00025233).

**Figure 1.**
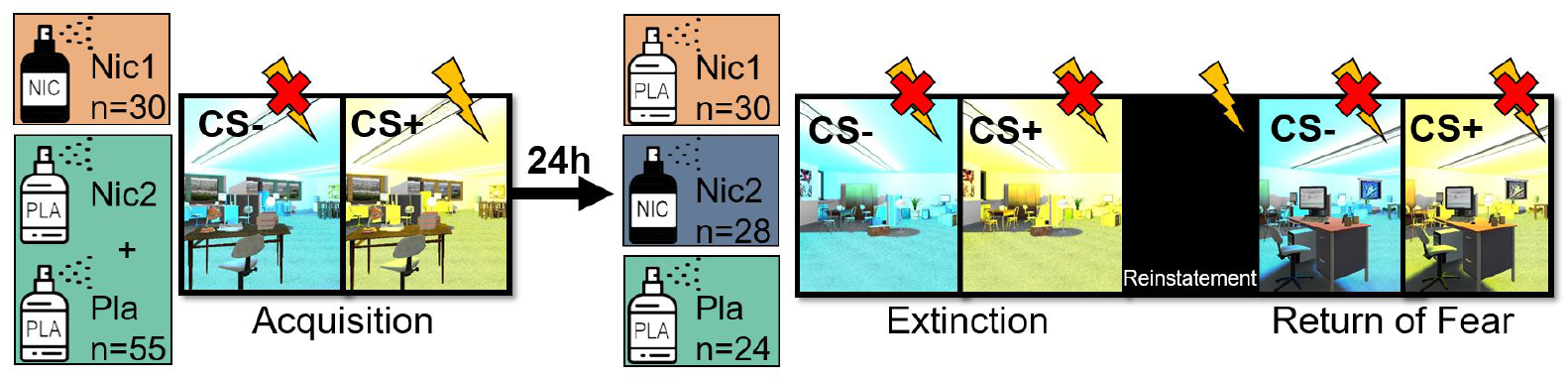
Experimental Procedure. The group Nic1 received nicotine before the Fear Acquisition on day 1 (ACQ), all remaining subjects received Placebo. After 24h the group Nic2 received nicotine and the remaining subjects received Placebo. Following that, subjects performed the Extinction training (EXT) with a subsequent Return of Fear (RoF) manipulation in form of a Reinstatement. CS colours were counterbalanced.

### Stimulus material

We used a two day context-dependent cue conditioning paradigm, which is an adaptation of an already described protocol^14^. The context was a virtual room that was presented on a screen (Source Engine, Valve Corporation, Bellevue, USA^15^). Three different contexts were used, where context A and context B were both virtual offices but differed in their set-up and context C was a mixture between context A and context B. Each context was presented from two different viewpoints. A cue was presented in each of these contexts, which was a coloured light that was illuminating the room in either yellow or blue (as described in^14^). One cue (CS+, duration of 6s) was predictive for an aversive electric stimulus (US, see below; 5.5s after CS+ onset), whereas another cue (CS-, duration of 6s) was not reinforced. Colours of the cues were counterbalanced. Context-presentations without cues were used as inter-trial intervals (ITIs, duration range between 7-11s). Visual stimuli were presented using Presentation® software (Version 20.3, Neurobehavioral Systems, Inc., Berkeley, CA, www.neurobs.com).

### Unconditioned stimulus

The electrotactile stimulus that served as a US consisted of a train of 3 pulses, each with a duration of 2ms and an interval of 50ms. The US was delivered via a surface electrode (Specialty Developments, Bexley, UK) on the right dorsal hand using a DS7A electrical stimulator (Digitimer, Welwyn Garden City, UK). The US intensity was individually adjusted prior to ACQ to a threshold that was perceived as very unpleasant, but not hurtful (mean=2.28mA, sd=4.09, min=0.4mA, max=37mA; no difference between groups, see below).

### Pharmacological intervention

Participants received 1mg nicotine as oral-spray (Nicorette® Spray, Johnson & Johnson GmbH) and 1mg placebo as oral-spray (St. Severin Cayenne Pepper Spray®, HECHT Pharma GmbH, 27432 Bremervörde), or placebo on both days. The dose was tested in a prior pilot study and proofed to be effective with little side effects. Typical side effects (dry mouth/skin, blurry vision, inertia, nausea, vertigo and headache) as well as self-reported side effects were documented after both experimental days. Participants rated their perception on a scale from 0 (no side effect) to 6 (extreme side effect). No difference of side effects was found between participants receiving nicotine and participants receiving a placebo (mean_Nic_=0.34, sd_Nic_=0.86; mean_Pla_=0.25, sd_Pla_=0.76).

### Outcome measures

#### Ratings

Participants rated how much fear/stress they felt towards the CSs and the ITIs on a visual Analogue Scale [VAS, 0 (none) – 100 (maximally)] before and after ACQ as well as before and after EXT and after the reinstatement test. Preceding the US presentation, subjects rated their US expectancy as a binary choice (yes/no) trialwise on both experimental days when the CSs were presented on the screen, (without any rating scale).

#### SCR

Skin conductance responses were measured on the hypothenar on the left hand of the participant and recorded with a BIOPAC MP-100 amplifier (BIOPAC® Systems Inc, Goleta, California, USA^15^). Raw data was scored manually. Criteria for a valid response were an occurrence between 1000ms and 4000ms after stimulus onset, a duration between 0.5s and 5s and an amplitude larger than 10nS. Null-reactions were defined as responses taking place later than 4000ms after stimulus onset or when there was no response. A missing response was defined as a response that starts before 1000ms after stimulus onset or if the responses were undefinable due to artifacts. Scored data was normalized for each day and participant (logarithmized and range-corrected).

#### Regions of interest

Regions of interest (ROI) were defined as key structures in emotional processing and fear, such as the bilateral amygdala, the bilateral hippocampus, bilateral insular cortex, dACC and vmPFC, and dopaminergic innervated key structures, such as the Nucleus Accumbens (NAcc) and the ventral tegmental area (SN/VTA) (definitions of ROIs in supplement).

#### Questionnaires

Participants gave information on age, gender, alcohol and coffee consumption, as well as their smoking behaviour. Furthermore, participants completed the State-Trait Anxiety Inventory (STAI-S/STAI-T)^16^. Questionnaires were filled out on Leiner.2019. To check if nicotine has a general effect on attention, participants completed the d2 Test of attention after each experimental day^17^ (for details see supplement).

### Data analysis

#### Rating/SCR

The analyses of the fear ratings (pre/post), the US expectancy ratings (trialwise), as well as the SCR (trialwise) were calculated using linear mixed effect models in R with the lme4 package^18^. As preregistered, we focused the analysis on the last block/post ratings of the ACQ and EXT. The dependent variable in the models were the ratings, or respectively the SCR.

Additionally, we implemented random intercepts for subjects. To test the paradigm, we analysed the interaction between stimuli-types (CS+/CS-) in the post Fear Ratings and during the last block for US expectancy/SCR and the groups (ACQ: nicotine (Nic1) / placebo (Nic2 and Pla); EXT: nicotine before day 1 (Nic1) / nicotine before day 2 (Nic2) / placebo (Pla)): RatingResults/SCR (1|participants)+stimulus*group). The Return of Fear analysis was performed similarly to the EXT, but we included the last three trials of the EXT, as well as the first three trials after reinstatement as blocks (model: RatingResults/SCR (1|participants)+stimulus*block*group (2 stimuli (CS+/CS-), 2 blocks (US expectancy and SCR: block 1: 3 trials before reinstatement, block 2: 3 trials after reinstatement; Fear Ratings: block 1: post EXT rating, block 2: post RoF rating), 3 groups (Nic1/Nic2/Pla)). To further investigate group effects across outcome measures, we used the same model, but the stimulus discrimination (CS+ - CS-) was calculated as dependent variable: StimulusDifference (1|participants)+group (additional *block in RoF analysis). To further test the results estimated marginal means (EMMs) were computed using the emmeans package as post-hoc tests and p-values were corrected for multiple comparisons using Bonferroni-Holms method. Regarding the differential post ACQ Fear Ratings, we used jasp^19^ to calculate an ANOVA with the dependent variable of the stimulus difference and calculated follow up post-hoc tests. This was necessary, because a linear mixed effects model with random intercept for each participant can only be fitted if more than one value of outcome measure per participant is included. This was not the case here. Additional analyses of the other blocks/pre ratings we performed accordingly.

#### fMRI

Preprocessing and statistical analysis of functional MRI data was carried out using SPM12 (Statistical Parametric Mapping, http://www.fil.ion.ucl.ac.uk/spm) running under MATLAB R2021b (The MathWorks, Inc., Natick, Massachusetts, United States). Before preprocessing, the first five volumes of each time series were discarded to account for T1 equilibrium effects. Remaining images were unwarped, realigned to the first image, coregistered to the individual high resolution T1 structural image, normalized (using DARTEL) and smoothed. Following statistical analyses were performed using a general linear model (GLM) at the single-subject level as standard approach for fMRI implemented in the SPM software. Experimental conditions (i.e., CS+, CS, (omitted) US, introductions, ratings, and button presses) were defined as separate regressors (as stick function) modelling the predicted time courses of experimentally induced brain activation changes. Subsequently a full-factorial analysis was calculated on the group level. As preregistered, we focused our analyses on the last block of ACQ and EXT, but added analyses of the other blocks. Additional analyses on both days were performed in which the US expectancy of each participant was used as parametric modulator (i.e., expectation of no US>expectation of a US). Participants that showed minimal variation in rated US expectancy (i.e., only one trial with no US expectancy) where removed from the analysis.

#### Connectivity analyses

To investigate functional connectivity differences in stimulus discrimination between groups, we employed psycho-physiological interactions (PPI, SPM12 standard approach) for the last block of the ACQ. As seed region served the left hippocampus as described in our ROIs. We then used the PPIs of each participant as regressor in an individual GLM, including movement as regressor. Finally, calculated estimates we contrasted on group level.

## Results

### Acquisition

Participants discriminated between CS+ and CSat the end of the Acquisition (ACQ) (last block/post Rating; pre-registered contrast) in Fear Ratings, US expectancy and SCR across all groups (for details see Supplement, Table S2/Figure S2-S4).

#### Group effects

In order to test if acute nicotine administration reduces CS-discrimination at the end of Fear Acquisition training, we compared CS-responses between the group that received nicotine (Nic1) with individuals that received placebo on day 1 (Pla). Our preregistered analysed focused on the last block during acquisition training (8 trials of each CS), based on our pilot data on US expectancy.

- Fear Rating. According to our hypothesis, we found lower differential Fear Ratings (CS+>CS-) after the ACQ in the group that received nicotine, when compared to the placebo group (preregistered post-hoc results: (t(165)=2.296, p_corr_=0.046; ANOVA: time*group interaction (F(1,83)=3.155, p=0.079); Figure 2a). This effect was driven by lower Fear Ratings towards the CS+ in the group that received nicotine, compared to the placebo group (t(83)=2.00, p=0.049; Figure 2a). There was no group effect in the analysis of the fear ratings before ACQ (F(1,83)=0.043, p=0.836), indicating that nicotine did not change baseline levels of subjective fear.
- US expectancy. We found no group effect on US expectancy in the last block of the ACQ (F(1,83)= 0.032, p=0.859; Figure S2).
- SCR. We found a stimulus*group interaction (F(1,928)=6.182, p=0.013, Figure S4), with higher differential skin conductance response in the group that received nicotine, when compared to the placebo group (t(60.1)=2.292, p_corr_=0.025; Figure S4). This enhancement in differentiation by nicotine was contrary to our pre-registered hypothesis and comparison of CS-specific responses between groups revealed no differences.
- fMRI. First, we aimed to delineate the neural effect of reduced CS-differentiation by nicotine which parallels the reduced CS-differentiation in subjective fear. This analysis revealed reduced differential (CS+>CS-) responses in the left (and trendwise in the right) hippocampus in the group that received nicotine, compared to the Placebo group (left HC: T=3.77, p_FWE_=0.016; right HC: T=3.36, p_FWE_=0.055; Table 1 and Figure 3). This effect was driven by a reduction in responses to the CS+ and thereby mirrored the reduced discrimination in rated fear in the group that received nicotine, compared to Placebo controls. The inverse contrast (i.e., enhanced discriminatory responses in the Nicotine, vs. the Placebo group) revealed no voxel within our ROIs. In order to establish the effect of nicotine on HC activation within the earlier stages of aversive learning, we compared the differential responses (CS+>CS-) between groups in the first and second block of ACQ, as an exploratory analysis. Similar, to the last block of ACQ, we found a similar decrease in the differential activation of the bilateral HC (left HC: T=3.60, p_FWE_=0.026; right HC: T=3.53, p_FWE_=0.033) in the group that received nicotine, compared to the placebo group in the first block (Table 1). In addition to this effect in the HC, we found decreased differential responses in the left AMY in the group that received nicotine, compared to the placebo group in the first block and trendwise in the second block of the ACQ (first block;left AMY: T=3.68, p_FWE_=0.009; second block; left AMY: T=3.03, p_FWE_=0.054; Table S2). We also found a similar effect in the left NAcc in the first block (T=3.03, p_FWE_=0.023). The adaptive differentiation between CS+ and CS-during aversive learning is reflected in the hippocampal (Figure 3), amygdala and NAcc (Figure 4) responses within the Placebo group, but is impaired after administration of nicotine.

**Table 1.**
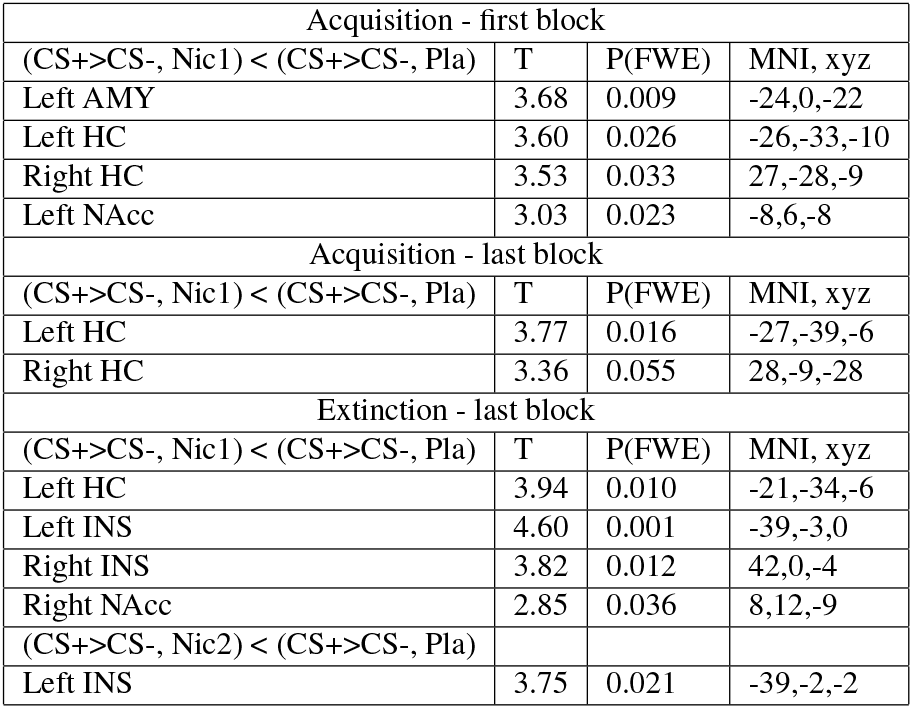
fMRI-results of ROIs (amygdala(AMY), hippocampus (HC), Ncl. Accumbens (NAcc), Insula (INS)). Group comparisons (Nic1=nicotine before Acquisition, Nic2=nicotine before Extinction, Pla=placebo group).

**Figure 2.**
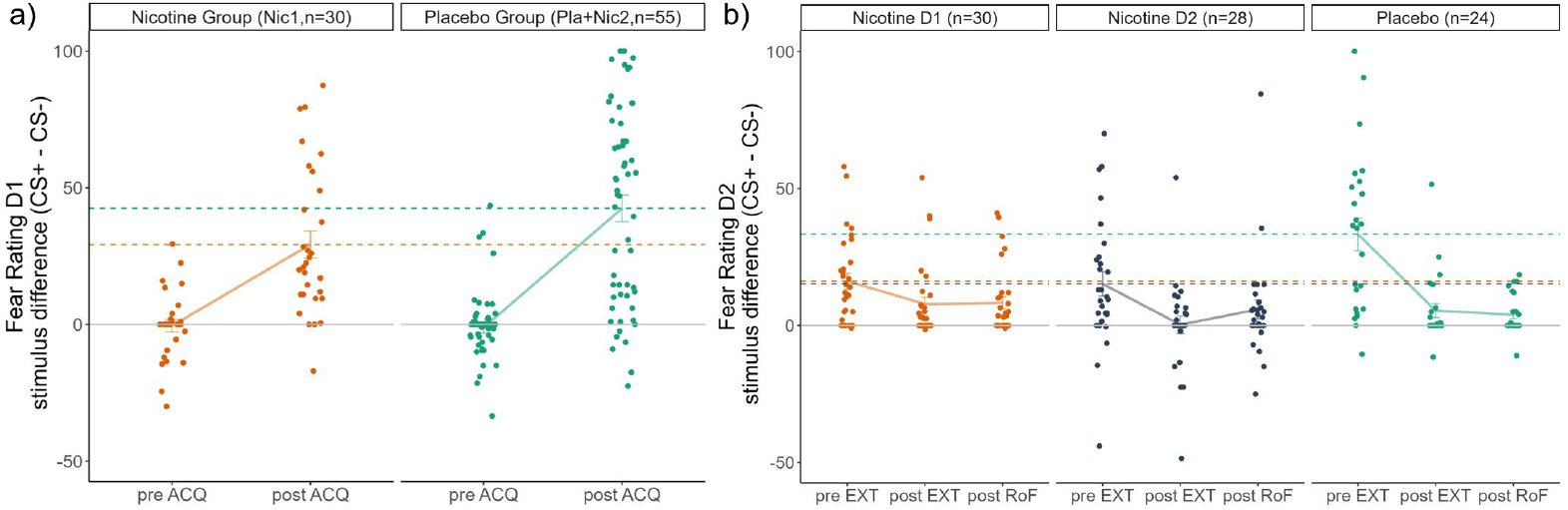
Weaker stimulus discrimination in the groups that received nicotine when compared to the Placebo group in Fear Ratings, a) after Fear Acquisition (day 1), b) before Extinction (day 2). No group differences in the differential Fear Ratings were found pre ACQ, post EXT or post RoF. Single subject responses are shown as scatterpoints, mean differential Fear ratings are depicted as lines with standard errors. Dashed lines represent the mean differential Fear Ratings per group post ACQ/pre EXT.

**Figure 3.**
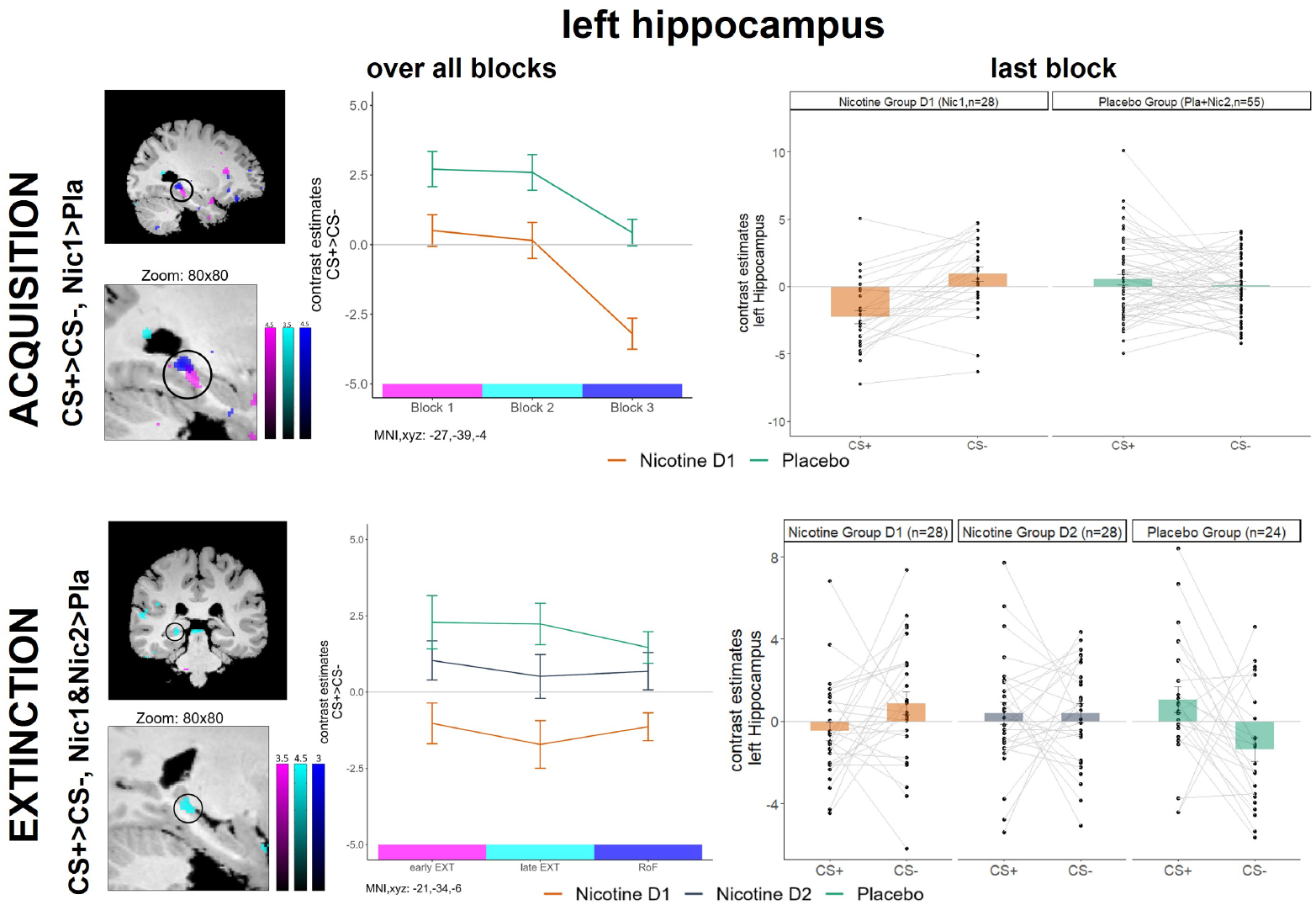
Acquisition: Acute nicotine administration, compared to placebo, reduces differential responses to the CS+ and the CS-during the last block of ACQ in the left hippocampus (similar to Fear Rating results). This effect is robust over all three blocks of the ACQ in the left HC. Extinction: Weaker stimulus discrimination during late EXT in the left HC in the group that received nicotine before ACQ (Nic1), when compared to placebo controls. No group differences in hippocampal activity during EXT or RoF were found between Nic2 and Pla. Scatterpoints represent single subject parameter estimates to each CS. Bars represent means across each group with standard error.

**Figure 4.**
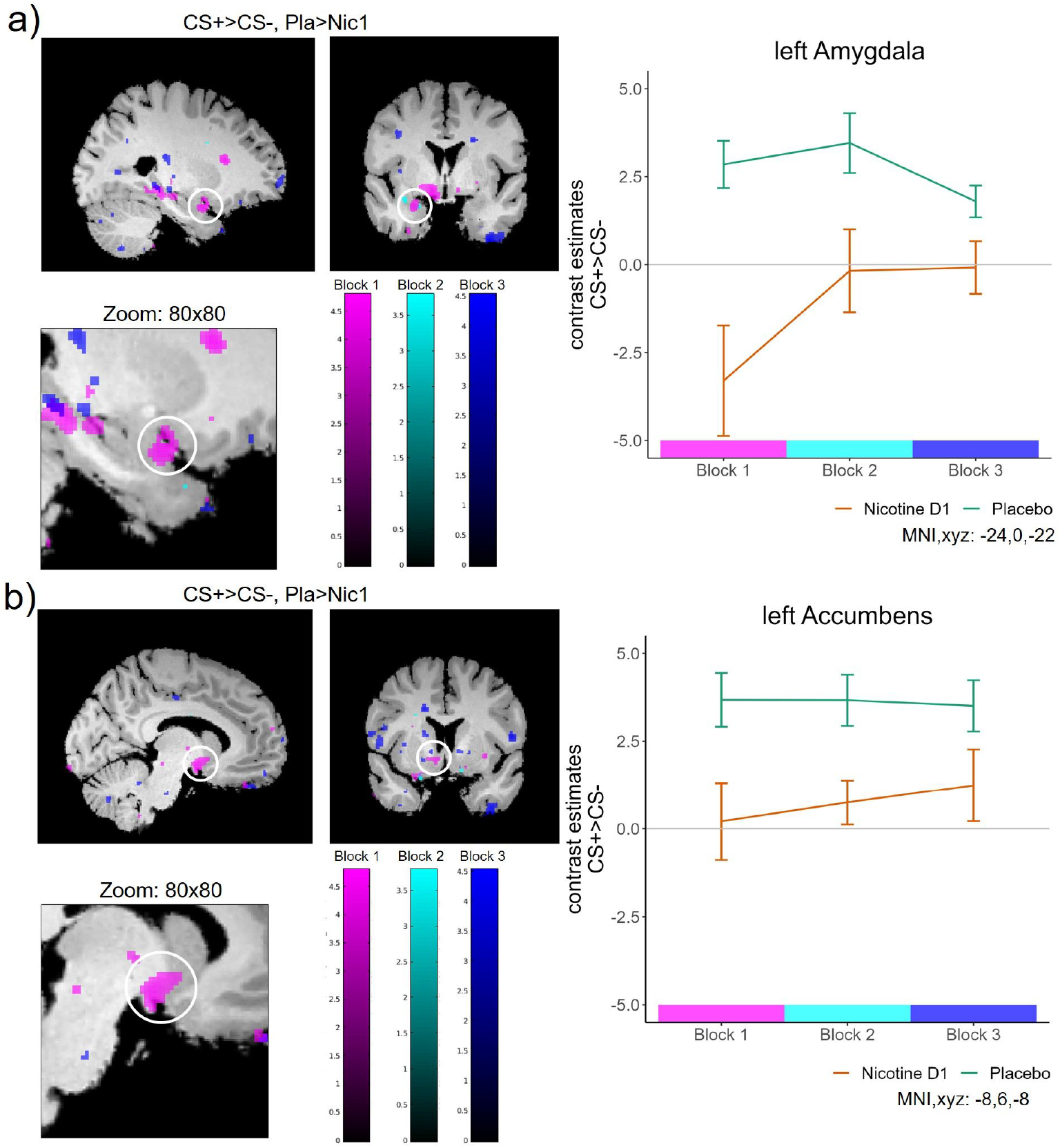
Stimulus discrimination during Fear Acquisition, fMRI results over all blocks in the a) left Amygdala and b) left Ncl. Accumbens. The group that received nicotine (Nic1) shows a lower activity in the ROIs over time, when compared to the Placebo group (contrast: CS+>CS-, Pla>Nic1). Parametric Modulations. In order to follow the individual learning of threat contingencies, we entered individual US expectancy ratings (if US expected: 1, if US not expected: -1) as a modulator for the activity towards the CS+. Hence, the activation in this analysis would reflect responses in a brain region that is modulated by US expectation during CS+ presentation (this analysis was similar to the preregistered analysis for the EXT, including the whole ACQ time-course). Here, we found that the activity in SN/VTA was modulated by US expectancy during CS+ presentation, which was altered by nicotine administration (MNI xyz: -10,-21,-12; T=4.80, p_FWE_<0.001; Figure 5). Specifically, we found activation in the placebo group in the SN/VTA when the US was not expected (see Figure 5 most right barplot). In contrast, nicotine administration led to activation of the SN/VTA when the US was expected (see Figure 5 most left barplot). In other words, the activity in the Placebo group decreased with increasing expectancy in the SN/VTA, whereas nicotine administration led to constant responses when participants expected a US.

**Figure 5.**
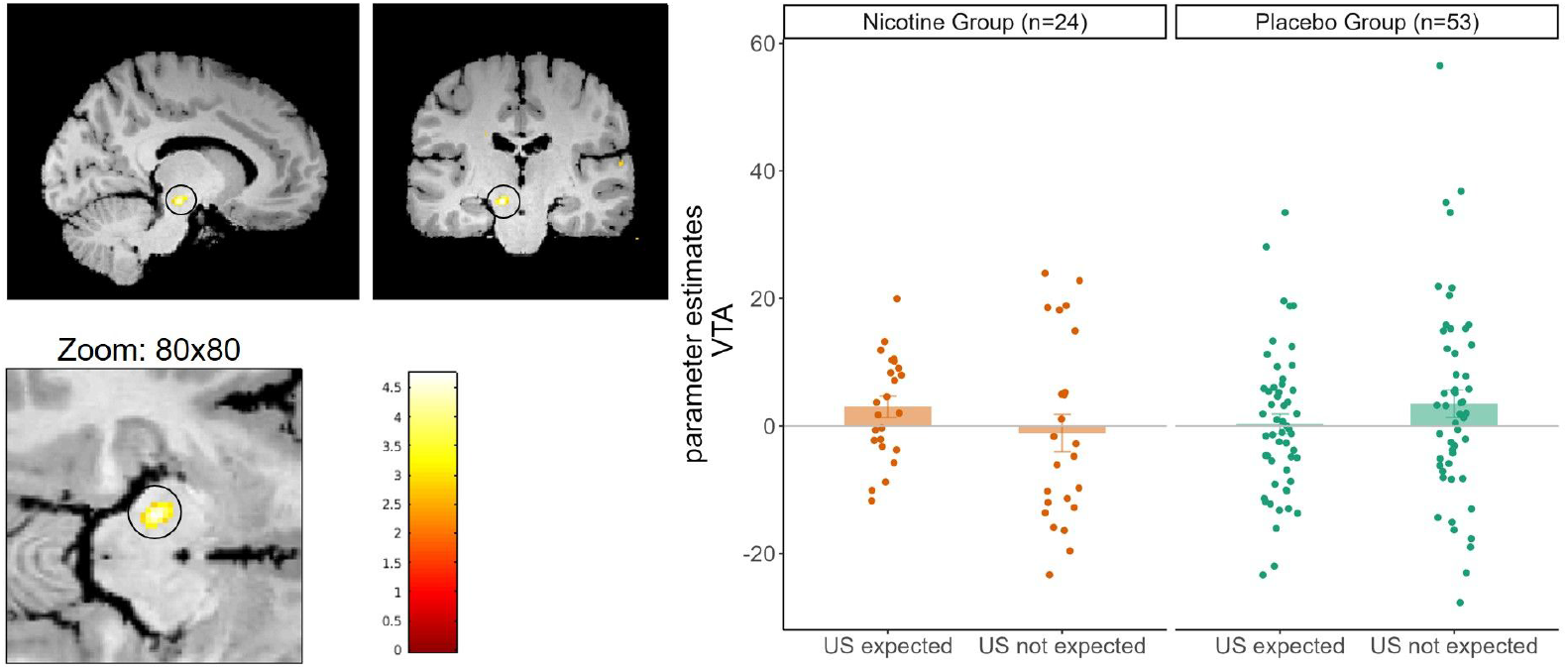
Stronger activation in the SN/VTA in the Nicotine group, when subjects expected the US, when compared to the Placebo group over the whole Fear Acquisition. Scatterpoints represent single subject contrast estimates in the SN/VTA, bars represent means with standard errors.
- PPI. To further investigate the linkage of the nicotine effect in the hippocampus and other ROIs, we employed a functional connectivity analysis in form of a psycho-physiological interaction (PPI). Our PPI analyses with the left hippocampus as a seed region was calculated for the last block of ACQ. We found increased functional connectivity in the group that received nicotine before ACQ between the left hippocampus and the bilateral AMY (left AMY: MNI xyz: -20,-6,-15; T=3.07, p_FWE_=0.064; right AMY: MNI xyz: 27,0,-14; T=3.45, p_FWE_=0.030) as well as the right Ncl. Accumbens (MNI xyz: 12,9,-10; T=2.82; p_FWE_=0.042), when compared to the Placebo group (Figure S1). An exploratory PPI for the whole ACQ confirmed these results.

### Extinction

On day 2, participants underwent Extinction training (EXT), in which CS-discrimination was still evident at the end of EXT across groups, indicated by higher Fear Ratings, US expectancy, trendwise higher SCRs and multiple ROIs (for details see Supplement, Table S3/Figure S2-S4).

#### Group effects

In order to examine if prior nicotine administration during acquisition training (Nic1), as well as acute nicotine administration during extinction (Nic2) resulted in diminished CS-discrimination, as compared to placebo (Pla), we compared responses between all three groups (Nic1/Nic2/Pla) in the last EXT block (i.e., eight trials; see preregistered analysis/methods).

- Fear Rating. Against our hypotheses, we found no effect of nicotine on the differential Fear Ratings at the end of EXT. Our secondary analyses compared CS-discrimination in subjective fear that was given before EXT, reflecting the retrieval of learned threat memories from day 1. Before EXT, we found a stimulus*group interaction (F(2,79)=4.949, p=0.009) with reduced differential Fear rating in both nicotine groups, when compared to the placebo group (Pla-Nic1: t(52)=2.734, p_corr_=0.009; Pla-Nic2: t(50)=2.472, p_corr_=0.017). Hence, these group differences suggest that the effect of reduced CS-differentiation by nicotine during learning endures to later (drug-free) retrieval. Furthermore, acute administration of nicotine also reduces CS-differentiation during retrieval, even when the learning was unaffected by the drug (Figure 2b). Additionally, our analysis revealed a main effect of acute nicotine administration on extinction of threat responses (F(2,124.48)=4.465, p=0.013), indicating trendwise increased Fear Ratings across both CSs in the group that received acute nicotine before extinction (Nic2), when compared to the placebo group (t(79)=2.237, p_corr_=0.084).
- US expectancy. We found a stimulus*group interaction (F(2,1219.09)=4.656, p=0.010, Figure S2) at the end of EXT, but post-hoc comparisons of CS specific responses between groups revealed no differences. Furthermore, we found no group effect on US expectancy in the memory retrieval during the first block of EXT (F(2,1213.15)=1.632,p= 0.196).
- SCR. The analysis of SCRs revealed a stimulus*group interaction (F(2,717)=4.922, p=0.008, Figure S4) in the last block of the EXT, which consisted of higher SCR towards the CS+ in the group of nicotine administration during acquisition (Nic1), when compared to the placebo group (Nic1 – Pla: t(70)=2.695, p_corr_=0.026; and compared to acute nicotine administration Nic1 – Nic2: t(70)=2.359, p_corr_=0.042). These results were against our hypothesis, but mirror the SCR results from day 1. We found no effects of nicotine administration on SCR during memory retrieval in the first block.
- fMRI. Next, we aimed to examine the effect of prior nicotine during acquisition (Nic1) and acute nicotine administration during extinction (Nic2) on hemodynamic responses during EXT (last block, as preregistered), both compared against placebo (Pla). Similar to our findings from ACQ, this analysis revealed that nicotine administration during acquisition of threat responses (Nic1), still led to reduced differential responses (CS+ > CS-) in the left HC during the last block of EXT (T=3.94, p_FWE_=0.010; Figure 3), when compared to the placebo group. This effect mirrors the effect of nicotine on patterns of hippocampal activity, as well as the Fear ratings, during acquisition of threat responses. Additionally, we found that nicotine during ACQ reduced differential responses in extinction training within the bilateral INS (left INS:T=4.60, p_FWE_=0.001; right INS: T=3.82, p_FWE_=0.012) and right NAcc (T=2.85, p_FWE_=0.036), when compared to the placebo group. The acute effect of nicotine on EXT was reflected by reduced differential responses in the left INS (T=3.75, p_FWE_=0.021; Table 1 and Figure S5), as compared to placebo controls. Exploratory analysis of the first block of the EXT, indicated a trendwise reduced discrimination after acute nicotine administration (Nic2), as compared to placebo in the left NAcc (T=2.67, p_FWE_=0.065). This effect mirrors the reduced discrimination in Fear ratings in this group.

## Discussion

Our study departed with the hypothesis that nicotine leads to decreased discrimination between danger and safety stimuli, when compared to the Placebo group. Our analyses of the Fear Ratings and fMRI results confirmed our hypotheses for both, acquisition and extinction of conditioned threats. During Fear Acquisition, we found a decreased discrimination of rated fear in the group that received nicotine, when compared to the placebo group. Reflecting the Fear Ratings, the nicotine group also showed a lower differential (CS+>CS-) hippocampal activation than the placebo group. This effect of nicotine administration on Fear Acquisition was further extended to a decreased discrimination in subjective fear between the CS+ and CS-during memory retrieval (before EXT) on day 2. Furthermore, the deficit in threat and safety discrimination in hippocampal activity by nicotine administration during ACQ was still evident during the end of EXT, when compared to the placebo group. In striking similarity to these effects, acute nicotine administration before EXT also resulted in decreased differential fear ratings between the CS+ and CS-, accompanied by decreased differentiation in Insular activity. The Return of Fear manipulation turned out to be relatively robust against nicotine effects over outcome measures. There is strong evidence from rodent and human studies that chronic nicotine disrupts discrimination of threat and safety learning^9,10,13^. Here we could show that already a single, small dose of acutely administered nicotine leads to similar changes in human learning processes. We further could show that nicotine alters hippocampal processes that enable discrimination between threat and safety. Such a neural effect by nicotine in humans is not only in line with animal studies in which nicotine enhanced synaptic plasticity in the hippocampus (LTP)^5,6^, but also with the role of the hippocampus in fear learning and extinction^20^. We further found that the functional connectivity between the hippocampus and the AMY and NAcc is enhanced by nicotine, which further aligns with our finding that both of these regions showed a similar decrement in discrimination of threat and safety signals during initial learning, as an effect of nicotine administration. Our results thereby imply that nicotine administration impairs differential activation to threat and safety cues within a network that is important for affective learning, including the hippocampus, AMY and NAcc.

A network-loop that includes the hippocampus and the NAcc has previously been described to process new salient information by integrating signals from the SN/VTA^21^. Furthermore, the SN/VTA in connection to the NAcc is part of the dopaminergic reward system, which is reinforced by nicotine intake^22^. In line with this, we found that nicotine led to activation of the SN/VTA towards the CS+, when participants expected the US, compared to placebo controls (importantly, the US expectancy did not differ between groups). The activation of the SN/VTA might be related to an amplification of a signal for a salient stimulus processing (CS+ when a US is expected). The impaired discrimination between threat and safety in the hippocampus, NAcc and AMY could be one source that results in overactivation of the VTA, by conveying enhanced salience (AMY) and novelty (hippocampus). Another possibility is that nicotine distorts VTA processing by ramping up phasic signals to a salient stimulus (CS+ when a US is expected), which results in distorted hippocampal learning and salience integration in the AMY. Both possibilities highlight that nicotine distorts salience processing (encoded in the SN/VTA), which is related to impaired threat discrimination (in fear ratings and neural activation in the hippocampus, NAcc and AMY). These finding would align with the finer grained animal models of salience and novelty encoding along a VTA-hippocampal loop^21^. Assuming this mechanism for nicotine during aversive learning in humans, it would mean that the integration of salient new information about CS-US contingencies into long-term memory is distorted. Such an impairment in processing salient information with the SN/VTA, hippocampus, NAcc and AMY may then underlie impaired discrimination between threat and safety. Such a reduced discrimination is thought to reflect clinically relevant maladaptive aversive learning, as the same effect is found in patients with Anxiety disorders (AD)^23^ and patients suffering from post-traumatic stress disorder (PTSD), showed decreased hippocampal activation during extinction recall, when compared to healthy controls^24^. Nicotine might effectively strengthen these deficits reported in patients with AD by distorting salience processing. The analysis of the US expectancy ratings, which rather reflects cognitive understanding of the paradigm, was not effected by nicotine on either day. Analyses of SCRs on the other hand revealed group effects that were contradictory to our hypotheses. We found a stronger stimulus discrimination in the group that received nicotine on day 1 during the last block in both ACQ and EXT than in the placebo group. As we find this effect only in the last block, nicotine does not simply increase skin conductance, but seems to lead to decreased habituation over time. These results were not reflected by any other outcome measures, but might reflect a previously reported effect, which could be a specific nicotinic influence on the sweat glands (which are innervated by acetylcholine)^25^.

The limitations of our study include that every participant regardless of their body weight received the same dose of nicotine (1mg). Additionally, we could only rely on self-reported non-smoking status, as well as drug and psychiatric disorder history. Furthermore, the placebo control group has a higher STAI-T score, when compared to both nicotine groups, which suggests a difference in trait anxiety. However, there was no difference between groups regarding their STAI-S score, which is a measure for the current anxiety state at the time point of the data collection. In fact, a higher STAI-T score would rather be associated with a weaker discrimination of threat and safety^26^, but we find the opposite effect in the Placebo group.

In conclusion, our study provided evidence how nicotine alters fear learning and extinction processes in humans. We found that nicotine impairs the discrimination between danger and safety. This is likely due to decreased differential activation in the hippocampus along with the amygdala and Ncl. Accumbens after nicotine administration. This study thereby reveals a mechanistic insight how nicotine administration leads to maladaptive aversive learning in humans and provides the neuropsychopharmacological link how smoking might contribute as a significant risk factor to the development and maintenance of pathological anxiety.

## Data Availability

The rating and SCR data, as well as the contrast fMR images used in this study are available for download.

https://gin.g-node.org/MadeleineMueller/Mueller_et_al_2023_Nicotine_and_fear

## Data availability

The rating and SCR data, as well as the contrast fMR images used in this study are available for download from: https://gin.g-node.org/MadeleineMueller/Mueller_et_al_2023_Nicotine_and_fear.

## Acknowledgements

The authors thank Katrin Bergholz, Kathrin Wendt, Waldemar Schwarz, Smilla Weisser and Jannis Petalas for help with MRI-data acquisition. We further thank Jürgen Finsterbusch for the development of the MRI sequence. This work was supported by the German Research Foundation (DFG) by an individual research Grant 7470/3-1 awarded to JH, and the SFB 936 (178316478 - C06) and the TRR 289 (422744262 - A06) awarded to JR.

## Author contributions statement

MM and JH conceived and designed the study paradigm. MM collected the data. JR and TF provided medical supervision. MM and JH analyzed the data. MM and JH drafted the initial manuscript, and TF and JR provided critical revisions. All of the authors revised the article and approved the final manuscript for submission.

