## Supplement for "Nicotine reduces discrimination between threat and safety in the hippocampus, nucleus accumbens and amygdala"

Material & Methods

Power analysis

A prior power analysis for a repeated-measures ANOVA (within and between subject interactions) for Fear Ratings indicated a sufficient sample size of 78 participants in total (26 per group) to detect an effect size=0,229 and a critical F=3.119 assuming a power (1-β error probability) of 0.95 and an α error probability of 0.05 (G*Power 3.1.9.4). Ten additional measurements of participants were conducted for the data acquisition, as the drop-out rate is quite high for pharmacological studies, as well as for fMRI studies. This concluded to a total of 88 participants after data acquisition.

Regions of interest

The amygdala, the hippocampus, the insular cortex and the NAcc were defined by Harvard-Oxford probability maps for the amygdala, the hippocampus, the insular cortex and the NAcc. For the dACC and vmPFC were no anatomical masks available, therefore we defined these two ROIs as in previous studies^1,2^. The vmPFC ROI was defined as a box of 20 × 16 × 16 mm at x=0 y=42 z=-12. The dACC ROI was defined as a box of 20 × 16 × 16 mm at x=0 y=28 z=26. The SN/VTA complex was defined by Bunzeck et al. ^3^. Correction for multiple comparisons within these ROIs was performed by using family-wise error correction based on the Gaussian Random Fields as implemented in SPM.

fMRI scan sequence

MRI data were obtained on a 3T Magnetom-PRISMA System (Siemens, Erlangen, Germany) using a 64-channel head coil. fMRI measurements were performed using single-shot echo-planar imaging with parallel imaging (GRAPPA, in-plane acceleration factor 2) and simultaneous multi-slice acquisitions ("multiband", slice acceleration factor 2). Echo planar multiband images were acquired with 42 continuous axial slices (1.5 mm thickness, 0.5mm gap) in aT2*-sensitive sequence (TR=1493ms, TE=30ms, flip angle=60°, field of view=225×225mm^2^). Moreover, high-resolution T1-weighted structural brain image (MP-RAGE sequence, 1mm isotropic voxel size, 240 slices) were obtained.

*Table S1: Demographics per group. Nic1 received nicotine before Fear Acquisition, Nic2 received nicotine before Extinction training and the control group received placebos before both days.*

|  | Nic1 | Nic2 | Pla |
| --- | --- | --- | --- |
|  | Mean (sd) | Mean (sd) | Mean (sd) |
| Sample size | 30 | 29 | 26 |
| Age [years] | 24.79 (4.39) | 26.17 (5.0) | 25.35 (3.98) |
| Gender | 56.67 % female | 55.17 % female | 55.56 % female |
| Coffee consumption [cups/day] | 0.89 (0.97) | 0.98 (1.22) | 0.7 (0.77) |
| Alcohol consumption [drinks/week] | 1.62 (2.07) | 0.83 (1.17) | 1.43 (1.43) |
| STAI-T | 40.2 (4.32) | 40.41 (4.47) | 43.31 (3.19) |
| STAI-S [day 1] | 42.9 (3.74) | 42.62 (3.97) | 42.58 (4.05) |
| STAI-S [day 2] | 43.13 (3.75) | 42.67 (4.83) | 43.2 (3.78) |
| Attention test - Day 1 [concentration performance score] | 252.43 (46.06) | 257.68 (38.88) | 266.74 (38.31) |
| Attention test - Day 2 [concentration performance score] | 272.03 (31.38) | 263.25 (37.89) | 273.24 (33.26) |
| Body Mass Index | 23.25 (2.70) | 23.67 (3.70) | 22.92 (3.17) |

Results

Acquisition

*Paradigm analysis last block of ACQ*

A stimulus discrimination after the ACQ was found in all three outcome measures. Specifically, we found a stimulus main effect in the Fear Ratings post ACQ (F(1,83)=23.65, p<0.001), where participants rated higher fear towards the CS+, compared to the CS- (t(83)=9.59, p_corr_<0.001). Similarly, analysis of US expectancy ratings during the last block revealed a stimulus main effect (F(1,1246.6)=334.18, p<0.001) with a higher US expectancy towards the CS+, when compared to the CS- (t(1247)=28.97, p_corr_<0.001). This stimulus effect was also found in SCRs during the last block (F(1,928)=12.049, p<0.001), with higher SCR towards the CS+ when compared to the CS- (t(928)=3.159, p_corr_=0.002). Analyses regarding the first/middle block can be found in the supplement.

Our predefined regions of interest (ROI) for fMRI-data analysis also revealed CS discrimination, indicated by higher hemodynamic responses to the CS+, as compared to the CS- during the last block of the ACQ within the bilateral Insula (left: MNI xyz: -32,20,4; T=9.23, p_FWE_<0.001; right: MNI xyz: 39,16,2; T=9.18, p_FWE_<0.001), dACC (MNI xyz: 3,22,27; T=6.72, p_FWE_<0.001), left NAcc (MNI xyz: -14,15,-6; T=3.43, p_FWE_=0.008), as well as in the SN/VTA (MNI xyz: 10,-22,-12; T=5.22, p_FWE_<0.001) across all groups.

*Analysis of the first and second block during ACQ*

Additional analysis included the examination of Fear Ratings, US expectancy and SCR towards the CS+, when compared to the CS- over all groups during the first and second block.

Regarding the **Fear Ratings**, we found no stimulus main effect pre ACQ (F(1,83)=0.036, p=0.851). There was no group effect in the analysis of the Fear Ratings before acquisition training (F(1,83)=0.043, p=0.836). The analyses of the **US expectancy** ratings revealed a main effect of stimulus in the first block (F(1,1228.48)=184.57, p<0.001), showing an increased US expectancy towards the CS+, when compared to the CS- (t(1227)=21.014, p_corr_ <0.001). The same effect was found in the middle block (F(1,1239.12)=397.92, p<0.001; CS+ > CS-: t(1239)=32.03, p_corr_ <0.001). No group effect was found in the first block (F(1,160.56)=0.001, p=0.974) or in the middle block (F(1,165.37)=0.035, p=0.851). The analyses of the **SCR** showed a stimulus main effect in the first block of the ACQ (F(1,928)=7.074, p=0.008), resulting from an increased SCR towards the CS+, when compared to the CS- (t(928)=3.964, p_corr_ <0.001). Regarding the middle block, we found a trendwise stimulus main effect (F(1,928)=3.049, p=0.081), again with an increased SCR towards the CS+, when compared to the CS- (t(928)=2.116, p_corr_=0.035). No group effects were found in the first block (F(1,115.21)=0.001, p=0.976) or in the middle block (F(1,94.8)=0, p=0.996).

Further analyses of the **fMRI** data were made regarding the differential contrast (CS+>CS-) between groups (Nic1< Pla) concerning the first and second block of the ACQ. Similar to the results from the analysis of the last block, we found reduced differential responses in the left HC (left HC: MNI xyz: -26,-33,-10; T=3.60, p_FWE_=0.026) and in the right HC (right HC: MNI xyz: 27,-28,-9; T=3.53, p_FWE_=0.033) in the first block. Additionally, we found reduced differential responses in the left AMY in the same contrast in the first block of the ACQ (left AMY: MNI xyz: -24,0,-22; T=3.68, p_FWE_=0.009), as well as trendwise during the middle block of the ACQ (left AMY: MNI xyz: -32,0,-18; T=3.03, p_FWE_=0.054). We also found an activation in the first block in the left Ncl. Accumbens (NAcc) for the same contrast (left NAcc: MNI xyz: -8,6,-8; T=3.03, p_FWE_=0.023). Additional main effect of group results of ROI activations during the ACQ can be found in table S2. Additionally, for the contrast (Nic1<Pla; main effect of group) during the middle block of the ACQ, we only found an activation in the right AMY (right AMY: MNI xyz: 32,-3,-21; T=3.17, p_FWE_=0.045). We furthermore compared responses across both CSs between groups within all three blocks. We found reduced responses in the right HC (MNI xyz: 24,-27,-12; T=3.69, p_FWE_=0.020), bilateral NAcc (left: MNI xyz: -6,14,-4; T=3.10, p_FWE_=0.020; right: MNI xyz: 12,12,-6; T=3.06, p_FWE_=0.018), SN/VTA (MNI xyz: 10,-24,-16; T=2.98, p_FWE_=0.068) and the bilateral AMY (left AMY: MNI xyz: -20,-4,-12; T=3.06, p_FWE_=0.052; right AMY: MNI xyz: 21,-9,-12; T=3.84, p_FWE_=0.007).

*Analysis of the main effects during the last block of ACQ*

Comparing **fMRI** data across both, the CS+ and CS-, we additionally found decreased activation in the nicotine, when compared to the Placebo group, within in the right HC (MNI xyz: 28,-30,-12; T=3.91, p_FWE_=0.011), bilateral NAcc (left: MNI xyz: -8,12,-6; T=3.18, p_FWE_=0.016, right: MNI xyz: 6,9,-6; T=3.47, p_FWE_=0.006) and SN/VTA (MNI xyz: 12,-22,-10; T=4.19, p_FWE_=0.002).

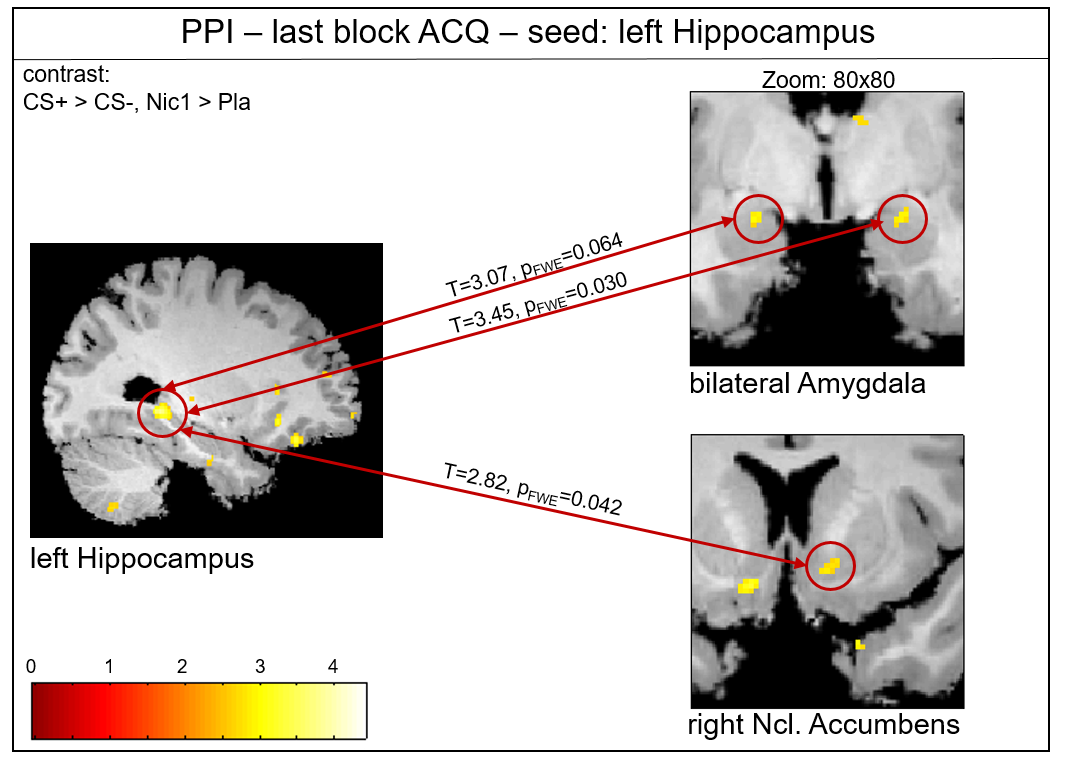

*Figure S1: Psycho-physiological interaction (PPI) during the last block of ACQ with left hippocampus as seed region. Nicotine administration before ACQ leads to increased connectivity towards the bilateral AMY and the right NAcc, compared to placebo controls.*

*Table S2: Fear Acquisition fMRI-results of ROIs. Group comparisons.*

| ACQ – first Block | Nic1<Pla | | |
| --- | --- | --- | --- |
|  | T | P(FWE) | MNI, xyz |
| Left AMY | 3.07 | 0.050 | -20,-4,-12 |
| Right AMY | 3.69 | 0.011 | 21,-9,-12 |
| Left HC | 3.94 | 0.009 | -28,-16,-18 |
| Right HC | 3.74 | 0.017 | 36,-24,-15 |
| Left INS | 4.08 | 0.006 | -38,0,0 |
| Right INS | 3.36 | 0.058 | 38,-2,-3 |
| dACC | 3.31 | 0.075 | -3,36,18 |
| Left NAcc | 3.05 | 0.022 | -8,15,-4 |
| Right NAcc | 3.06 | 0.018 | 12,12,-6 |
| SN/VTA | 3.04 | 0.058 | 9,-26,-18 |
| ACQ – middle Block | Nic1<Pla | | |
|  | T | P(FWE) | MNI, xyz |
| Right AMY | 3.17 | 0.045 | 32,-3,-21 |
|  | (CS+>CS-, Nic1) < (CS+>CS-, Pla) | | |
| Left AMY | 3.03 | 0.054 | -32,0,-18 |
| ACQ – last Block | CS+>CS- | | |
|  | T | P(FWE) | MNI, xyz |
| Left INS | 9.23 | <0.001 | -32,20,4 |
| Right INS | 9.18 | <0.001 | 39,16,2 |
| dACC | 6.72 | <0.001 | 3,22,27 |
| Left NAcc | 3.43 | 0.008 | -14,15,-6 |
| SN/VTA | 5.22 | <0.001 | 10,-22,-12 |

Extinction

*Paradigm analysis last block EXT*

On day 2, participants underwent Extinction training (EXT), in which CS-discrimination was still evident at the end of EXT across groups, indicated by higher Fear Ratings, US expectancy and trendwise higher SCRs (Fear Rating post EXT: stimulus effect F(1,79)=8.795, p=0.004; CS+>CS-: t(79)=2.811, p_corr_ =0.006; US expectancy last Block EXT: (F(1,1219)=84.176, p<0.001; CS+>CS-: t(1219)=12.203, p_corr_<0.001; SCRs last Block EXT: F(1,717)=3.065, p=0.080, CS+>CS-: no difference in post-hoc tests; for results of the first block, see Supplement) . The differential CS responses were furthermore reflected in the fMRI data, were we found increased activation in multiple ROIs in the differential stimulus contrast CS+>CS-, including the vmPFC, (Table S3) across all groups.

2

*Table S3: Extinction training (last Block) fMRI-results of ROIs of the whole sample.*

| EXT – last block | **CS+>CS-** | | |
| --- | --- | --- | --- |
|  | T | P(FWE) | MNI, xyz |
| Left HC | 3.30 | 0.070 | -32,-15,-21 |
| Left INS | 6.44 | <0.001 | -42,12,0 |
| Right INS | 4.98 | <0.001 | 34,22,-3 |
| dACC | 4.01 | 0.012 | -3,21,24 |
| vmPFC | 4.24 | 0.006 | 8,50,-20 |
| SN/VTA | 3.03 | 0.067 | -8,-24,-14 |

*Analysis of the first block of EXT*

Further examinations of our paradigm included the discrimination between CS+ and CS- during Extinction training, we examined Fear Ratings, US expectancy and SCR towards the CS+, when compared to the CS- over all groups during the first block/pre Rating.

Before the first stimulus presentation we found a stimulus main effect in the pre EXT **Fear Rating** (F(1,79)=14.783, p<0.001). Post-hoc tests revealed that participants rated higher fear towards the CS+, when compared to the CS- (t(79)=8.427, p_corr_<0.001). There was no main effect of group before the EXT (F(2,135.44)=2.168, p=0.118). Stimulus by group interactions are described in the main manuscript. Regarding the **US expectancy**, we found a stimulus main effect over all groups during the first block of the EXT (F(1,1212.08)=180.112, p<0.001). This effect stems from increased US expectancy towards the CS+, when compared to the CS- (t(1213)=21.853, p_corr_<0.001). There was no group effect in the first block of the EXT (F(2,118.62)=1.352, p=0.263). No main effects of stimulus (F(1,717)=0.751, p=0.386) or group (F(2,73.61)=0.705, p=0.497) during the first block was found in the **SCR**.

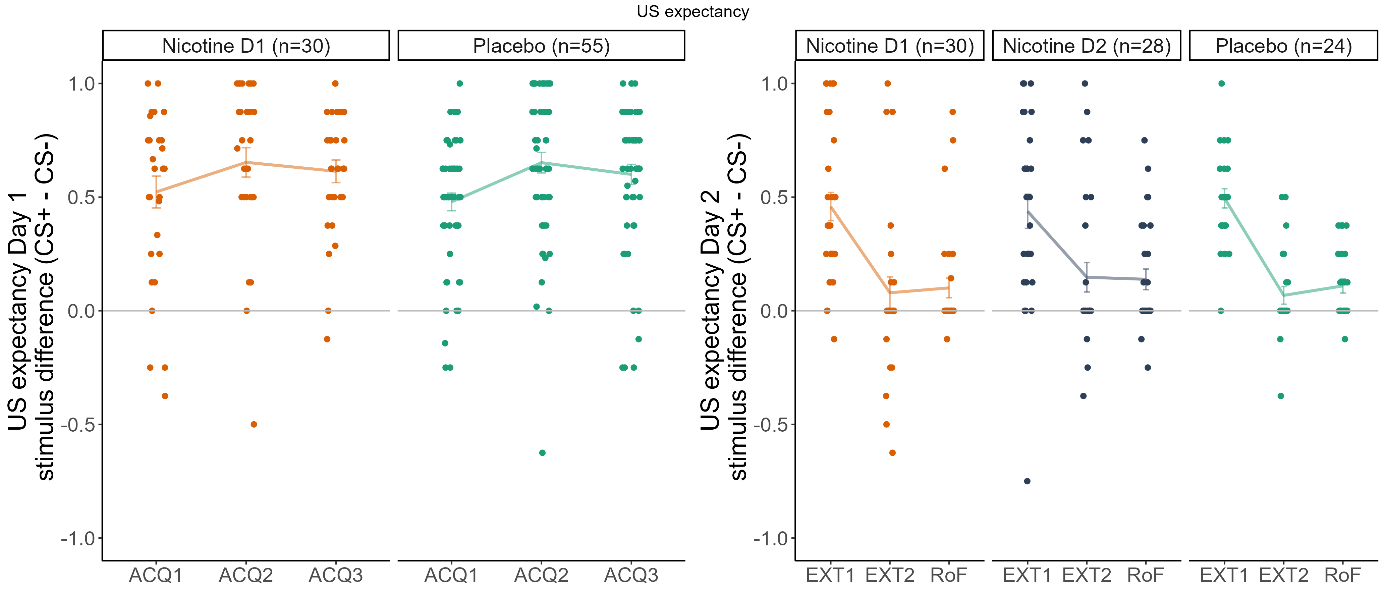

*Figure S2: No group differences were found a) during the last ACQ3 or b) EXT2 in the US expectancy.*

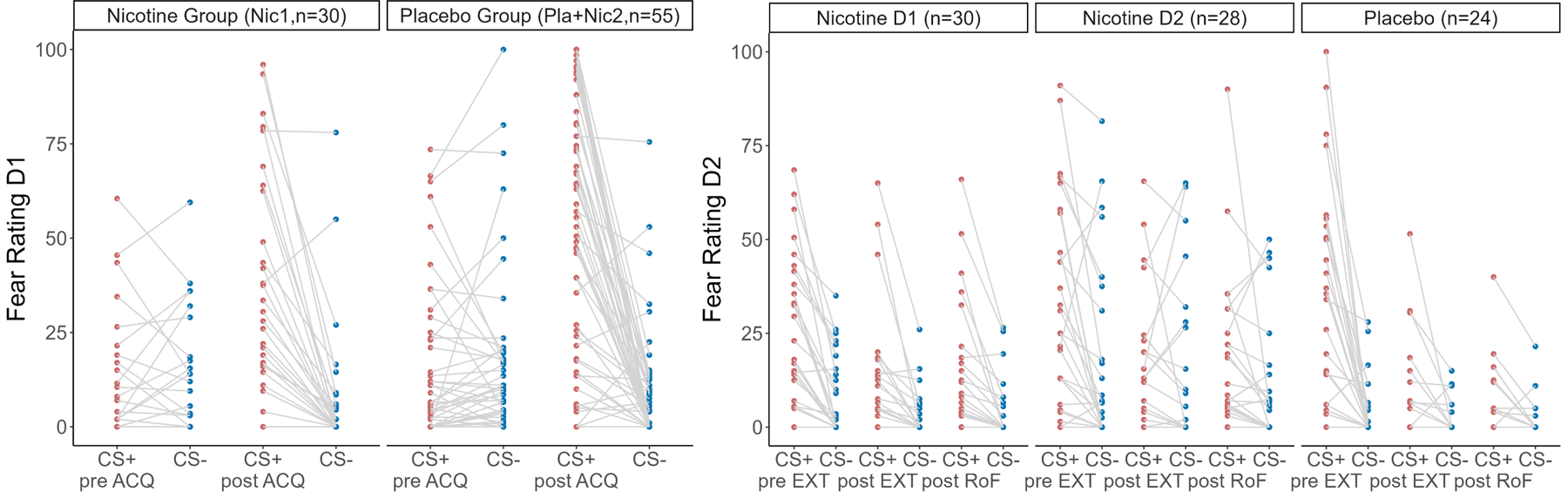

*Figure S3: Nicotine (administered during ACQ or during EXT) weakened discrimination between CS+ and CS- in Fear Ratings. Single subject responses are shown as scatterpoints.*

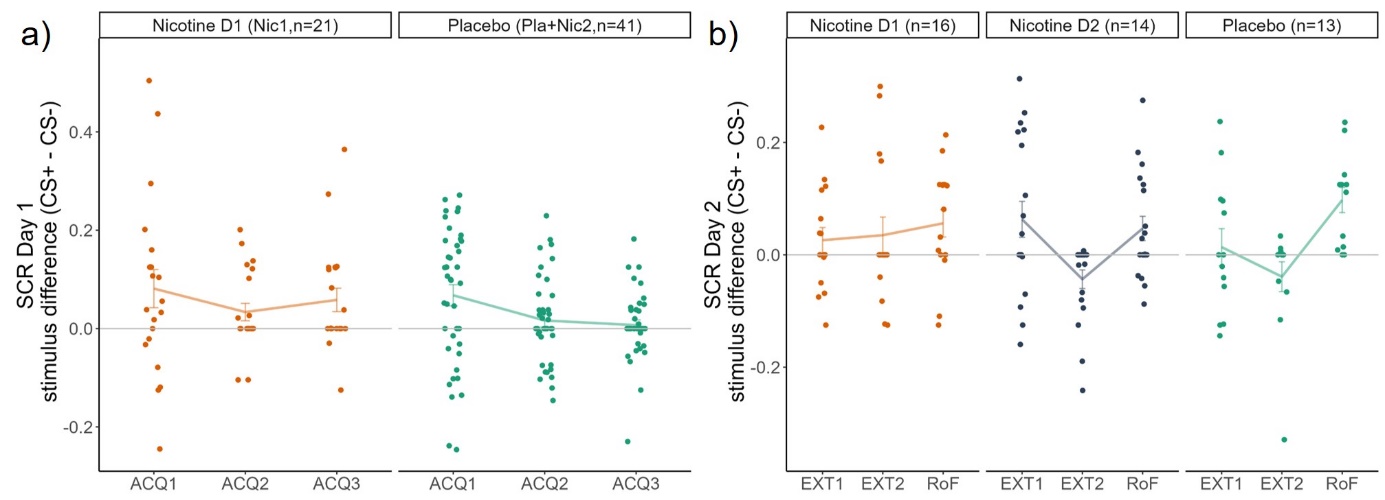

*Figure S4: Differential Skin conductance Responses a) during day 1 and b) during day 2. a) During the last block the differential SCR is higher in the Nicotine group, when compared to the Placebo group. b) During EXT2 the group that received nicotine on day 1 showed a trendwise increased SCR, when compared to the Nicotine group on day 2. There were no group effects concerning RoF.*

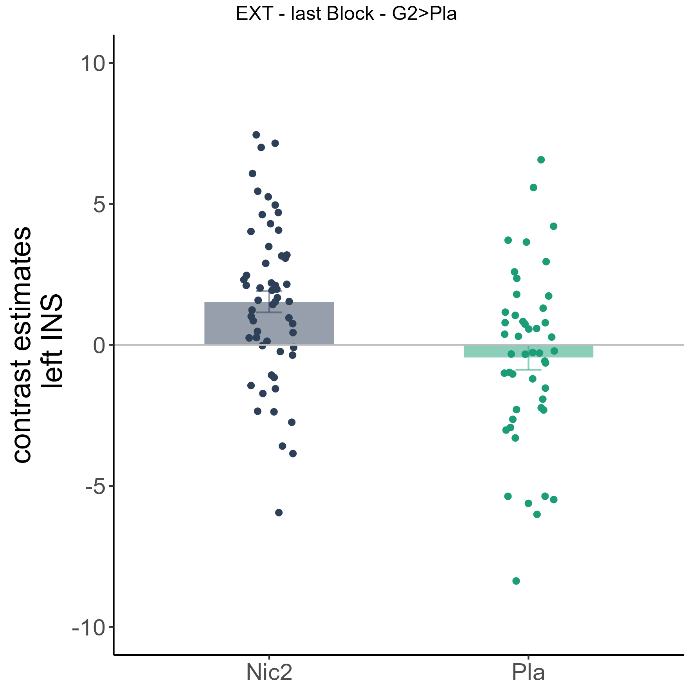

*Figure S5: The Nicotine group D2 showed increased activity in the left INS, when compared to the Placebo group during the last block of the EXT. Bars represent means across each group with standard error.*

*Analysis of the main effects during the last block of EXT*

Furthermore, exploratory **fMRI** analysis across both CSs during the last block of extinction revealed increased activity in the group that received nicotine during acquisition (nic1) in the left AMY (MNI xyz: -30,0,-22; T=3.40, p_FWE_=0.024), as well as trendwise in the right HC (MNI xyz: 27,-14,-20; T=3.27, p_FWE_=0.078), when compared to placebo controls. Within the opposite contrast (Nic1<Pla), we found enhanced activity in the vmPFC (MNI xyz: 8,42,-20; T=4.05, p_FWE_=0.010) and a trendwise in the dACC (MNI xyz: -6,33,28; T=3.44, p_FWE_=0.062) for the placebo group, when compared to nic1. The same analysis of acute effects of nicotine on extinction training revealed increased neural activity in the left INS (MNI xyz: -36,-10,9; T=3.65, p_FWE_=0.029; see figure S5), which is in line with increased Fear ratings in nic2.

Return of Fear

*Paradigm analysis RoF*

The Return of Fear was employed in this experiment by an application of reinstatement US after extinction training, which was followed by CS presentation in a context that was a mixture of the acquisition and extinction context (as reported in Esser et al. ^4^). We expected an increase of Fear Ratings, US expectancy and SCR following the reinstatement across groups.

The analysis of paradigm confirmed that US expectancy increased after reinstatement, when compared to before reinstatement. US expectancy analyses indicated a stimulus by block interaction (F(1,877.29)=3.895, p=0.049). We found that both stimuli showed an increase in US expectancy after the reinstatement (CS+, pre-post reinstatement: t(878)=6.363, p_corr_<0.001; CS-, pre-post reinstatement: t(878)=2.049, p_corr_=0.041). Comparing the stimulus discrimination before and after reinstatement (CS+>CS-, RoF>EXT), we found a trendwise effect in the SCR analysis, but no effect in Fear Ratings or fMRI.

*Group effects*

Fear Rating

The differential stimulus analysis investigating group effects revealed only an effect in the Fear Ratings. Here, we found a block by group interaction (F(2,79)=3.558, p=0.033), which consists of an increase in differential Fear Ratings in Nic2 after the RoF compared to after the EXT (t(79)=2.945, p_corr_=0.013). When testing which stimulus was crucial for this effect, we found that the Fear Ratings after the RoF in Nic2 towards the CS+ are higher, compared to the CS- (t(237)=5.857, p_corr_=0.044). Nic2 reported higher subjective fear towards the CS+ after RoF, when compared to the placebo group (t(162)=2.577, p_corr_=0.022). There was no effect in Nic1 (t(79)=0.255, p_corr_=0.8) or Pla (t(79)=-0.742, p_corr_=0.921).

fMRI

Regarding only the RoF block, we found increased differential activation in the group that received nicotine before ACQ (Nic1), when compared to the Placebo group in the vmPFC (MNI xyz: 9,50,-10; T=2.68, p_FWE_=0.004). This was driven by increased activation towards the CS- in the Placebo group (see Figure S6).

Combining the last block of the EXT and the RoF block, we found increased activation in the right NAcc in Nic2, when compared to Nic1 (MNI xyz: 10,-10,-10; T=2.84, p_FWE_=0.033) and furthermore trendwise increased activation in the bilateral INS (left INS: MNI xyz: -33,18,-2; T=3.35, p_FWE_=0.058; right INS: MNI xyz: 42,-6,6; T=3.30, p_FWE_=0.067). Additionally, we found a trendwise increased activation in the last EXT block and the RoF block in Nic2, when compared to Pla in the left INS (MNI xyz: -36,-10,10; T=3.17, p_FWE_=0.094). Furthermore we found trends towards an increased discrimination (CS+>CS-) in the RoF block in Nic2, when compared to Nic1 (right AMY: MNI xyz: 30,-6,-18; T=2.91, p_FWE_=0.089; MNI xyz: 30,-14,-20; T=3.13, p_FWE_=0.097). These data can be found in table S4.

When comparing the discrimination between (CS+>CS-) in the last block of EXT to the Reinstatement Test, we found increased activations in the left HC (MNI xyz: 34,-21,-18; T=3.87, p_FWE_=0.010), right INS: (MNI xyz: 38,-12,14; T=3.46, p_FWE_=0.043), dACC (MNI xyz: -3,21,24; T=3.36, p_FWE_=0.069) and vmPFC ( MNI xyz: 4,34,-20; T=3.75, p_FWE_=0.021, see table S4).

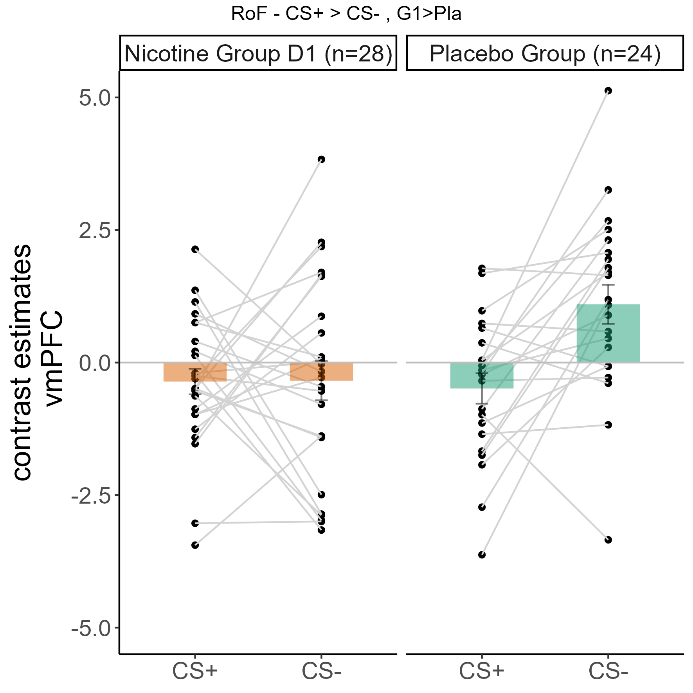

*Figure S6: The Nicotine group D1 showed increased differential activity in the vmPFC compared to the Placebo group during the RoF, because of increased activation towards the CS- in the Placebo group. Bars represent means across each group with standard error.*

*Table S4: Return of Fear fMRI-results of ROIs. Group comparisons.*

| RoF | Nic1<Nic2 (RoF+EXT) | | |
| --- | --- | --- | --- |
|  | T | P(FWE) | MNI, xyz |
| Left INS | 3.35 | 0.058 | -33,18,-2 |
| Right INS | 3.30 | 0.067 | 42,-6,6 |
| Right NAcc | 2.84 | 0.033 | 10,-10,-10 |
|  | Pla < Nic2 (RoF+EXT) | | |
| Left INS | 3.17 | 0.094 | -36,-10,10 |
|  | (CS+>CS-, Nic2) < (CS+>CS-, Nic1), (RoF) | | |
| Right AMY | 2.91 | 0.089 | 30,-6,-18 |
| Right HC | 3.13 | 0.097 | 30,-14,-20 |
|  | CS+>CS-, RoF<EXT | | |
| Left HC | 3.87 | 0.010 | 34,-21,-18 |
| Right INS | 3.46 | 0.043 | 38,-12,14 |
| dACC | 3.36 | 0.069 | -3,21,24 |
| vmPFC | 3.75 | 0.021 | 4,34,-20 |

Other analyses

*Finite Impulse responses*

To ensure that vasoconstrictive effects of the nicotine did not interfere with our analysis of hemodynamic brain responses, we calculated the activity in different ROIs over a time period of 10sec (10 time bins; Figure S7). The activity did not differ between groups.

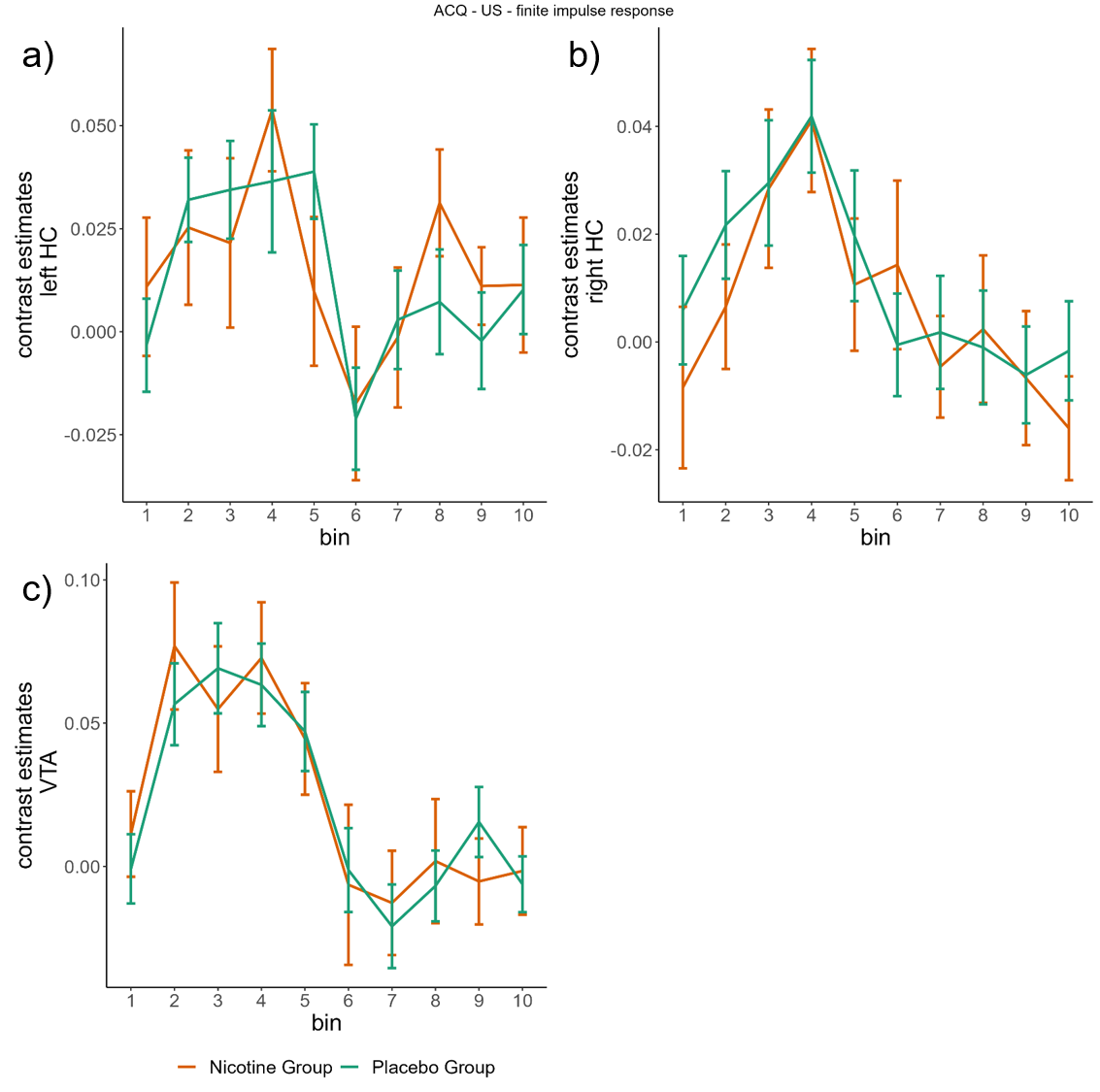

*Figure S7a-c: Finite impulse response. The activity pattern of both groups on day 1 over the time of 10sec after US presentation in the bilateral HC (a) and b)) and c) the SN/VTA.*

*D2 – Test of attention*

To test the general effect of nicotine on attention, subjects underwent a test of attention on both days. A rmANOVA revealed a main effect of day (F(1,5989.33)=27.207, p<0.001). Post-hoc tests showed that subjects achieved better results on day 2 (t(84)=5.199, p_corr_<0.001), likely because it was the second time they underwent the test and already had a routine. Furthermore we found a trend towards a day*group interaction (F(2,1304.57)=2.963, p=0.057), but post-hoc tests showed no difference.

To check the nicotine influence of nicotine for each day individually, we performed an independent samples t-test comparing the results of the attention test between the group that received nicotine on that respective day and the other two placebo groups (day 1: Nic1 vs Pla; day 2: Nic2 vs Nic1&Pla) and found no effect of nicotine on the attention of the subjects (day 1: t(84)=-1.02, p_corr_=0.313; day 2: t(84)=-1.148, p_corr_=0.254).
